# Bridging the gaps: Contextualizing the mhGAP Humanitarian Intervention Guide to implement in Pakistan

**DOI:** 10.1101/2025.05.21.25327990

**Authors:** Asma Humayun, Noor ul Ain Muneeb, Arooj Najmussaqib, Israr ul Haq, Muhammad Asif

**Affiliations:** Mental Health Strategic Planning and Coordination Unit Ministry of Planning, Development & Special Initiatives; KRL Hospital Islamabad; Ministry of Planning, Development & Special Initiatives Islamabad, Pakistan

**Keywords:** mhGAP-HIG, contextualization, cultural adaptation, Pakistan, LMICs, implementation gaps, primary healthcare, mental health and psychosocial support, MHPSS

## Abstract

**Background:** The mhGAP Humanitarian Intervention Guide (mhGAP-HIG) offers an evidence-based intervention to strengthen the capacity of non-specialists to manage mental, neurological, and substance use conditions in humanitarian contexts. The Ministry of Planning, Development and Special Initiatives (MoPD&SI) developed a Mental Health and Psychosocial Support (MHPSS) service model for Pakistan in 2021. As part of the model, this study identified implementation gaps and described the process of contextualizing mhGAP-HIG to address them.

**Methods:** We explored the implementation gaps through a three-step process, including a desk review, a focus group discussion, and four key informant interviews with relevant stakeholders. A thematic analysis using the framework method was applied. We followed the World Health Organization’s recommended process for contextualization and conducted an adaptation workshop. This was followed by a comprehensive review of the guide with reference to the country’s healthcare context and clinical practices, and a Delphi consensus achieving 90% success. We field tested the contextualized guide with training of trainers; and training of primary healthcare workers using pre- and post- knowledge assessment, analyzed via paired t-test.

**Results:** We identified four implementation gaps: knowledge, skills and attitude, knowledge-to-practice, training resource, and supervision gaps. While contextualizing, these gaps were addressed by incorporating additional knowledge; cultural constructs; instructions for interview, examination and interventions; and translating these instructions into Urdu language. To address the training resource gaps, the MoPD&SI published the adapted guide as a national resource; and developed a mobile application that serves as a complete reference and provides integrated features for remote supervision. The field testing of the adapted guide demonstrated its educational effectiveness, relevance, user-friendliness, and feasibility for implementation.

**Conclusions:** This study presents a systematic contextualization and field-testing of mhGAP-HIG. It demonstrates that successful contextualization requires a culturally grounded, system-aware, and stakeholder-informed approach. We recommend integrating the guide into pre-service training and ensure policy support for integration into primary care for long-term outcomes.

## Introduction

The World Health Organization’s (WHO) Mental Health Gap Action Programme (mhGAP), an evidence-based initiative aimed at reducing the treatment gap and scale up mental healthcare by enhancing the abilities of non-specialists, has been implemented in many countries across the world (R. C. Keynejad et al., 2018; World Health Organization, 2016, 2023). However, it remains challenging to implement the mhGAP Intervention Guide (mhGAP-IG) in some resource-limited settings, where only partial implementation has been feasible (Bitta et al., 2020a; R. Keynejad et al., 2021a; Li et al., 2015; Searle et al., 2022; Zheng et al., 2025). Barriers to scale up mhGAP interventions are well-documented, even in high-resource countries, such as stigmatization, financing, multi-stakeholder collaboration, legal and policy structures, accessibility, competencies of providers, and inadequate quality control, including supervision and referral systems (Woodward et al., 2023). The WHO strongly recommends that the mhGAP interventions be adapted to the local context to overcome prevailing challenges (World Health Organization, 2016).

In humanitarian contexts, where resources are even more constrained, the implementation of the mhGAP-IG presents additional challenges. Moreover, the mhGAP-IG does not explicitly cover stress and trauma-related conditions that are particularly prevalent in emergency settings (Echeverri et al., 2018; Ventevogel et al., 2015). To address this gap, the mhGAP Humanitarian Intervention Guide (mhGAP-HIG) was developed as a concise, practical, and easy-to-use tool for resource-limited settings, especially in the humanitarian context (World Health Organisation & UNHCR, 2015). Adaptation of the mhGAP-HIG is also essential to respond to the implementation challenges; to address culture-specific contexts, especially in humanitarian settings, to integrate local idioms of distress, coping methods, and approaches to care (Gómez-Carrillo et al., 2020). Some countries have implemented the mhGAP-HIG without adaptation and reported the barriers to implementation (Echeverri et al., 2018; Tarannum et al., 2019). For instance, Tarannum et al. (2019) acknowledged that health system preparedness, support from decision-makers to scale up, and clinical supervision are crucial for effective implementation of the guidelines. They also identified the language barrier as a limitation in their intervention. In some cases, a hybrid approach utilizing content from both mhGAP-IG and mhGAP-HIG has also been used. For example, in Bangladesh, mhGAP-IG was adapted for the humanitarian situation by incorporating ‘Acute Stress’ and ‘PTSD’ modules from mhGAP-HIG for the training sessions (Momotaz et al., 2019). They attempted to address some challenges by incorporating healthcare and cultural context into role plays and delivering the training in the local language. Ukraine has also followed a similar blended model of mhGAP for capacity building through the inclusion of stress-specific conditions from the mhGAP-HIG modules (PTSD, Acute Stress, and Grief) (World Health Organization, 2021). However, the adaptation processes were not documented.

A significant part of Pakistan has been facing severe challenges due to a prolonged humanitarian crisis driven by geo-political instability, violence, terrorism, the impact of climate change, hosting a large refugee population, and economic difficulties (European Commission, 2024; Harris et al., 2020; International Organization for Migration, 2024; UNICEF, 2023; Yaqoob et al., 2023). The need for mental health and psychosocial support (MHPSS) services in the country gained recognition after the earthquake in 2005 (M. M. Khan, 2006; World Health Organization, 2005). Subsequent initiatives were primarily short-term emergency responses, often lacked coordination and sustainable funding (Humayun et al., 2016; Riaz et al., 2023; UNICEF, 2020).

The process of exploring existing challenges, attempts to adapt the mhGAP-HIG in the healthcare and cultural context, and initiatives to build the capacity of non-specialists in Pakistan extends across many years (Humayun et al., 2016, 2017). Earlier field experiences indicated challenges, such as resource shortages: insufficient primary care staff, absence of community services, unavailability of psychotropic medications; biomedical approach of medical practice; and stigma surrounding mental disorders (Humayun et al., 2017). The need to incorporate stress and trauma-related conditions in training primary care physicians (PCPs) had also been highlighted (Humayun et al., 2016). While certain challenges have been identified during field experiences, further exploration is needed, particularly in relation to the mhGAP-HIG guidelines, by involving all relevant stakeholders in the process.

It is important to note that current mental health services depend primarily on psychiatrists, while clinical psychologists (CPs) represent an underused and inadequately trained resource. The WHO also endorses mhGAP interventions be incorporated in the training of PCPs and other allied health professionals from pre-service level onwards to improve mental health outcomes over the long term (World Health Organization, 2025).

In addition, the COVID-19 pandemic further highlighted the country’s substantial mental health burden and scarcity of specialist services. As a response, the Ministry of Planning, Development & Special Initiatives (MoPD&SI) developed a scalable, rights-based MHPSS model featuring a digital solution for MHPSS service provisions and centralized data management (Humayun, 2021; MoPDSI, 2022b). One of the objectives of this model is to address resource gaps, with a focus to strengthen capacity through training primary healthcare workers (PHCWs) in mhGAP-HIG.

This paper identifies implementation gaps, describes the process of contextualizing the mhGAP-HIG, and field testing of the adapted guide.

## Method

This study was carried out as part of the MHPSS project initiated by the MoPD&SI (letter no. 6(262) HPC/2020). Our primary objectives were to:

a. Explore the implementation gaps in the context of Pakistan.
b. Document the process of contextualizing the mhGAP-HIG to bridge the gaps.
c. Field test the training resources (mhGAP-HIG-PK guide and mobile application).

### 1. The implementation gaps

To explore the implementation gaps, we completed a three-step process: 1) a desk review 2) Focus Group Discussion (FGD) and 3) Key Informant Interviews (KIIs).

To gather and analyze existing information; and to understand mental healthcare needs and limitations of existing resources, we performed a desk review of publicly available data. Our data sources included the Ministry of National Health Services, Regulations and Coordination (MoNHSR&C) official website, published peer-reviewed articles, and reports from local and international organizations (Dayani et al., 2024; Humayun et al., 2016, 2017; S. A. Khan & Ahmed Khan, 2019; MoNHSR&C, 2019; World Health Organisation, 2021; World Health Organization, 2024).

Subsequently, we conducted a FGD to outline implementation gaps and engaged a diverse group of stakeholders, including PCPs (8), health administrators (3), staff from humanitarian agencies (iNGOs) (5), and mental health experts (8). A trained moderator facilitated the discussion using a semi-structured interview guide *(See supplementary material 1)*.

In addition, we conducted four KIIs with senior psychiatrists with extensive experience in teaching PCPs and were part of earlier mhGAP trainings in Pakistan. All participants of the FGD and KIIs were approached using snowball sampling. Prior to participation, verbal consent was obtained from all participants, and discussions were audio-recorded for accuracy.

We encoded the collected data and performed thematic analyses using the framework approach (Gale et al., 2013). Two independent researchers transcribed the interviews and re-checked the content of the transcripts with audio recordings. Subsequently, we used an inductive approach to manually assign initial codes. A third researcher collaborated with the initial coders to review the codes and consolidate them into categories. We organized the categorized data into a spreadsheet and conducted charting to summarize findings across all transcripts. Through a series of team discussions and iterative reviews, the data were interpreted to ensure accuracy and consistency.

### 2. Contextualizing the mhGAP-HIG to bridge the gaps

The contextualization process aimed at addressing the following implementation gaps whilst maintaining alignment with the mhGAP-HIG guidelines:

2.1 Knowledge, skills and attitude gaps

2.2 Knowledge to practice gaps

2.3 Training resources gaps

2.4 Supervision gaps

We followed a multiphase approach to contextualize the guide in accordance with the recommended mhGAP adaptation process (World Health Organization, 2016). Each gap was addressed through distinctly defined steps. For the first gap, we conducted an adaptation workshop including four mental health specialists, seven PCPs, and four health administrators. Following this, a team of three specialists—including two psychiatrists and a clinical researcher— systematically reviewed the guide with reference to the country’s existing health policies, clinical protocols, and guidelines used in pre-service training of PCPs. Table 2 presents the identified gaps and proposed changes.

For the knowledge to practice gaps, we added evidence-based interview questions for assessment, instructions for examination, and delivering psychosocial interventions.

These interview questions and instructions were translated into Urdu through the following process.

A bilingual team fluent in English and Urdu, comprising a psychiatrist and a psychologist, was assigned the task to translate the original English draft (M1), collaboratively working to synthesize their individual translations, into a cohesive Urdu version (M2). This step ensured the preservation of grammatical, semantic, and linguistic integrity. Subsequently, another team of two mental health experts fluent in both languages undertook the back-translation of M2 into English. This approach allowed for a direct comparison between the back-translated English version and the original English draft (M1). Through this direct comparison, the team was able to promptly identify and address any deviations in grammar, meaning, and language usage, thus streamlining the translation process.

A panel of six senior mental health experts were selected from across the country based on their extensive experience in teaching, delivering, and supervising mental health services in both community and humanitarian conflict settings. They evaluated the adapted draft using a Delphi approach (Jorm, 2015); and provided critical feedback for further refinement. The content of the guide was finalized after 90% consensus was reached (Diamond et al., 2014).

To address the third gap related to the training resources, the adapted draft (mhGAP-HIG-PK) was printed and a mobile application, mhGAP-HIG-PK was developed, which provided full access to the adapted guide in digital format. Both resources (mhGAP-HIG-PK guide and mobile application) were developed for training and future reference during clinical practice. To address the supervision gaps, additional features were integrated into the mhGAP-HIG-PK mobile application to enable trained participants to seek remote supervision.

### 3. Field testing the training resources

The final step of the study involved field testing the training resources to evaluate their feasibility and effectiveness. These activities were conducted online due to COVID-19 pandemic.

a. Training of trainers (ToTs)

Ten experienced trainers (eight psychiatrists and two clinical psychologists) were selected for a 5-day online ToTs through convenience sampling. Inclusion criteria of these trainers were at least 5 years clinical experience, 3 years teaching or training experience, and motivation and willingness to provide six months of remote supervision. Subsequently, these trainers delivered a 5-day training course, covering one module each. After the training, each trainer compiled a comprehensive report, as feedback, on their respective modules, identifying its strengths and weaknesses.

a. Training of primary healthcare workers (PHCWs)

For the second component, using purposive sampling, 13 PHCWs, including 5 PCPs with at least two years of clinical experience and 8 CPs – fresh graduates of postgraduate clinical psychology programs, were recruited. Their knowledge was assessed online using the mhGAP-HIG knowledge questionnaire (World Health Organization, 2022). Both pre- and post- scores were analyzed using a paired sample t-test. All participants provided their qualitative written feedback upon completion of the training.

Following field testing and the collection of feedback from both trainers and PHCWs, minor adjustments were made to the final draft of the guide. Subsequently, the MoPD&SI published the adapted guide as a national resource in 2022.

## Results

This section presents identification of implementation gaps, including a situational analysis of the contextual landscape of Pakistan, outcomes of the process of contextualizing the mhGAP-HIG, and field testing the training resources.

### 1. Implementation gaps

We present an overview of Pakistan’s mental health care needs and existing resources. The humanitarian situation in Pakistan has remained precarious over the last decade, with many regions experiencing multiple forms of conflict ranging from terrorism and violence to geopolitical disputes, and is consistently ranked among the ten most vulnerable countries to the effects of global climate change (Yaqoob et al., 2023). The effects of climate change make the country especially vulnerable due to its high rates of multidimensional poverty (Harris et al., 2020); and hosts one of the largest displaced populations (over 3.6 million Afghan refugees) in the world (European Commission, 2024). There has been a call to action for policymakers in Pakistan to improve mental healthcare delivery and incorporate MHPSS into disaster risk reduction strategies (UNDRR, 2019). Following the floods in 2022, an estimated 20.6 million people continued to need humanitarian assistance in 2024 (International Organization for Migration, 2023). In the humanitarian context, it is estimated that at least 1 in 5 people exposed to the above crises will require MHPSS services (Charlson et al., 2019).

Even beyond the humanitarian context, the ongoing burden is expected to be substantial. The evidence-based data inputs from Global Burden of Disease (2019) estimated that depressive disorders were the second leading cause of all-age Years lived with Disability in Pakistan (Hafeez et al., 2023). A recent systematic review outlined the factors that hinder access to mental health services, which include financial and logistics constraints, stigma, and lack of encouragement from family, reduced mental health literacy, dissatisfaction from previous treatments, and reliance on religious leaders and faith healers (Choudhry et al., 2023). The government spends a minuscule percentage of its health expenditure on mental health, which is estimated to cover less than 2% of the total economic burden of mental disorders (Alvi et al., 2023).

As for the general healthcare context, the devolution of power to provinces in 2010 aimed to help allocate increased resources towards health, planning, and the regulation of healthcare delivery in Pakistan. Still, effective translation remains impeded by weak institutional capacity and federal-provincial coordination (S. J. Khan et al., 2023; Zaidi et al., 2019). Consequently, there is a massive shortage of hospitals, doctors, paramedic staff, and most life-saving medications (Muhammad et al., 2023). A survey showed that more than half of the flood-affected districts did not have a single psychiatrist (Humayun, 2022). Additionally, there is a critical brain drain of highly qualified medical practitioners, attributed to suboptimal remuneration, hectic work schedules, job insecurity, and non-recognition of services (Meo & Sultan, 2023; Nadir et al., 2023). The medical practice, in general, is not person-centered as doctors are not trained to practice a collaborative approach to invest in informing people or seeking their preferences for treatment options (Jalil et al., 2017).

Table 1 presents our findings under four implementation gaps:

**Table 1:**
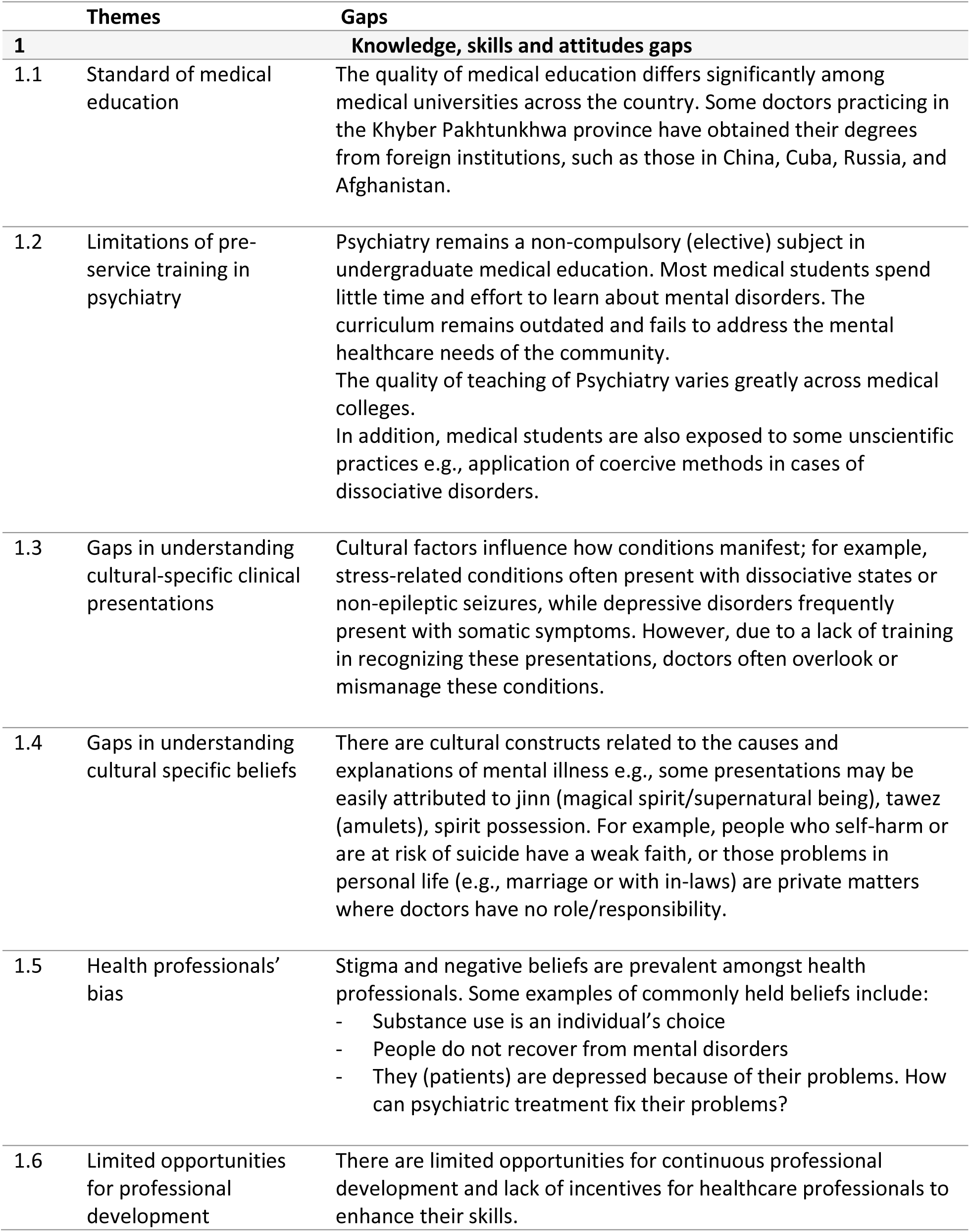
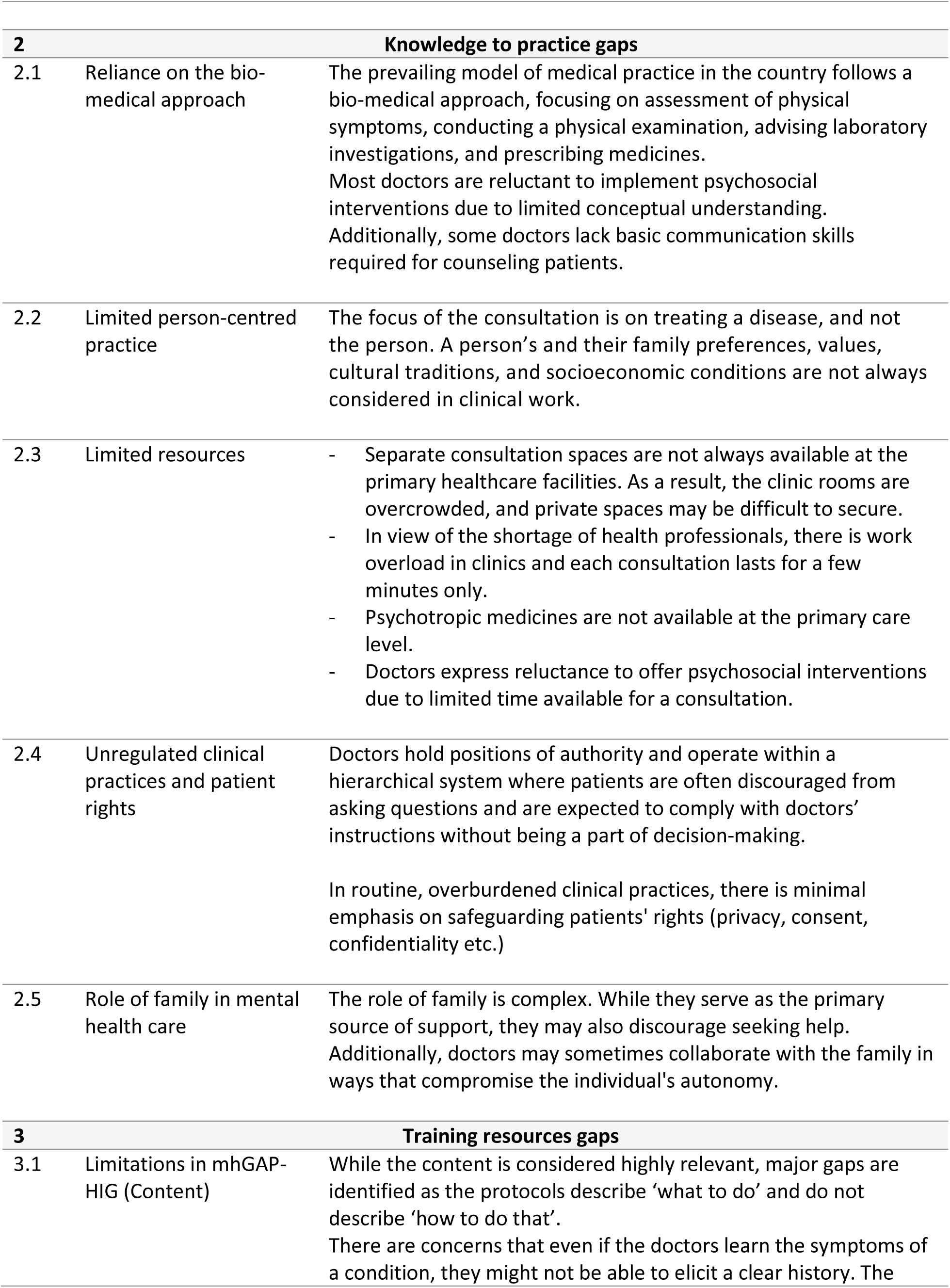
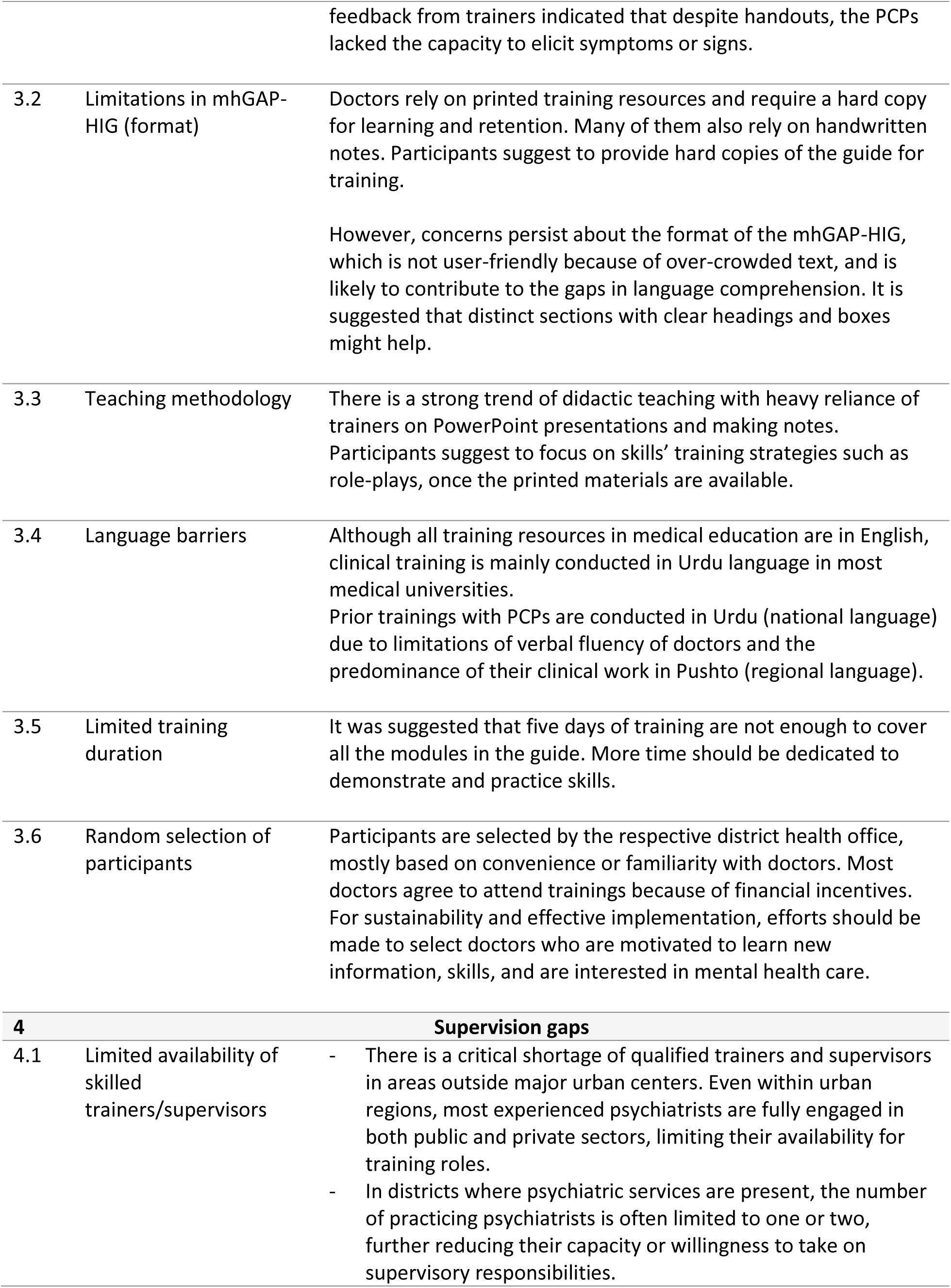
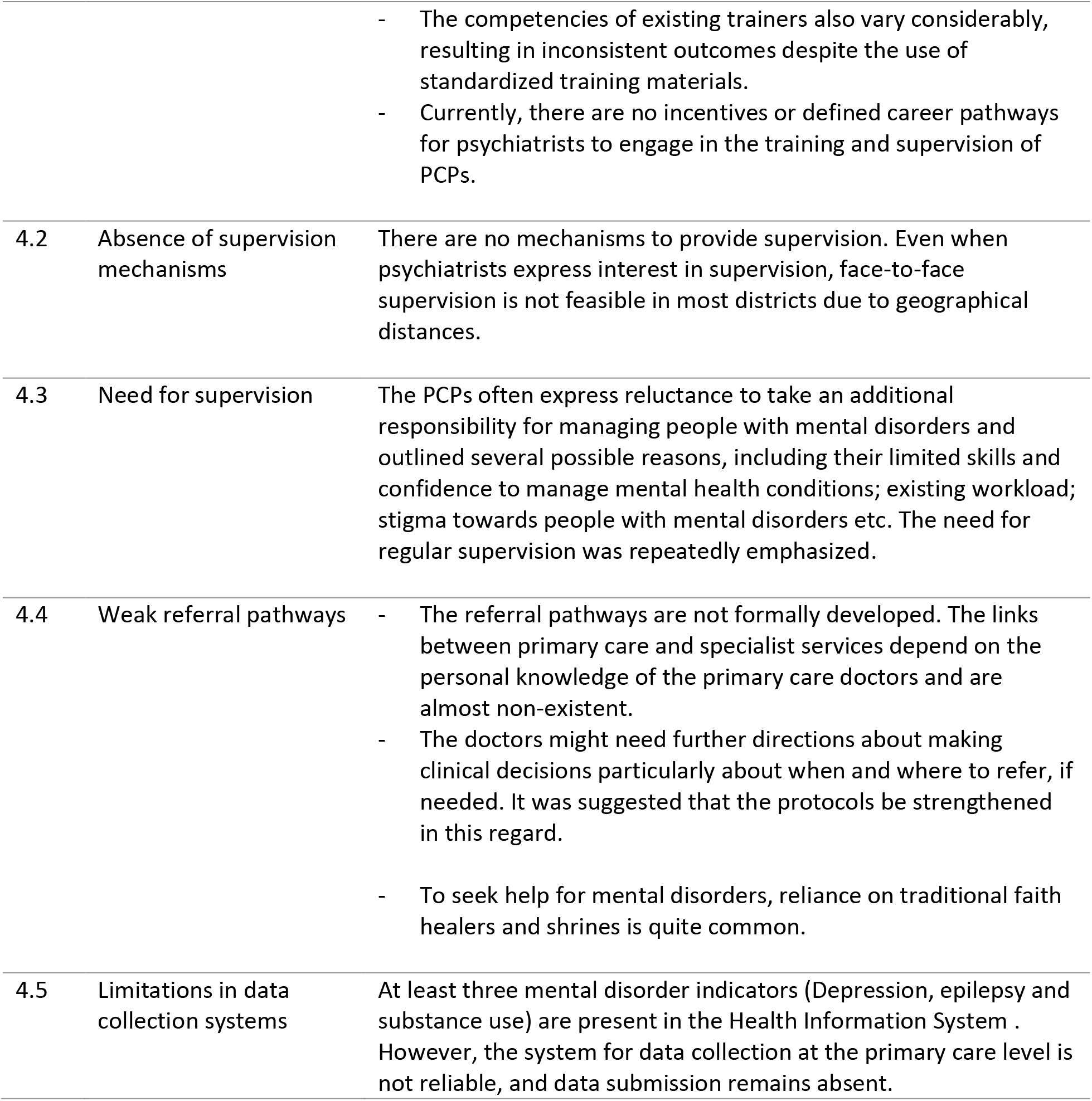
Implementation gaps.

**Table 2:**
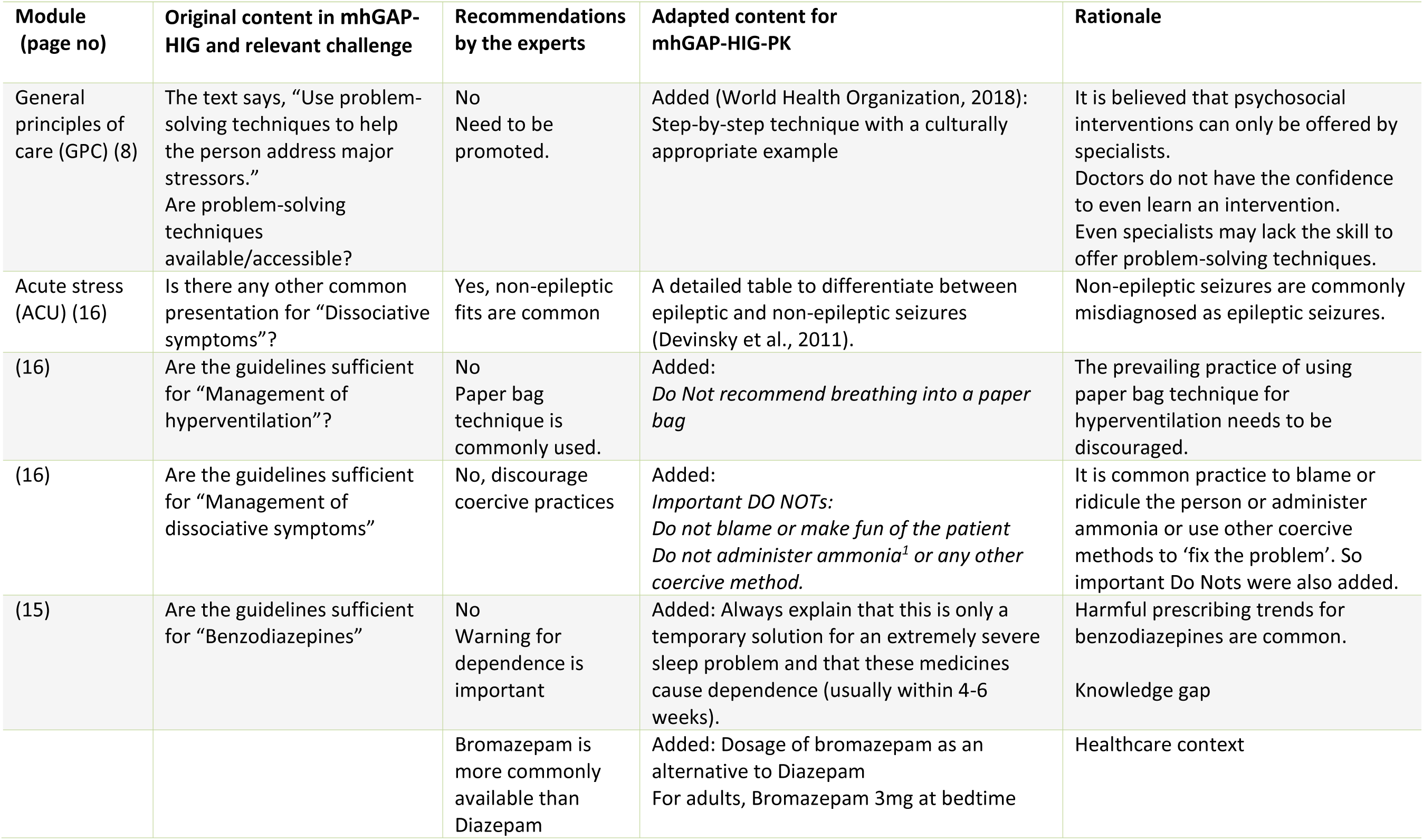
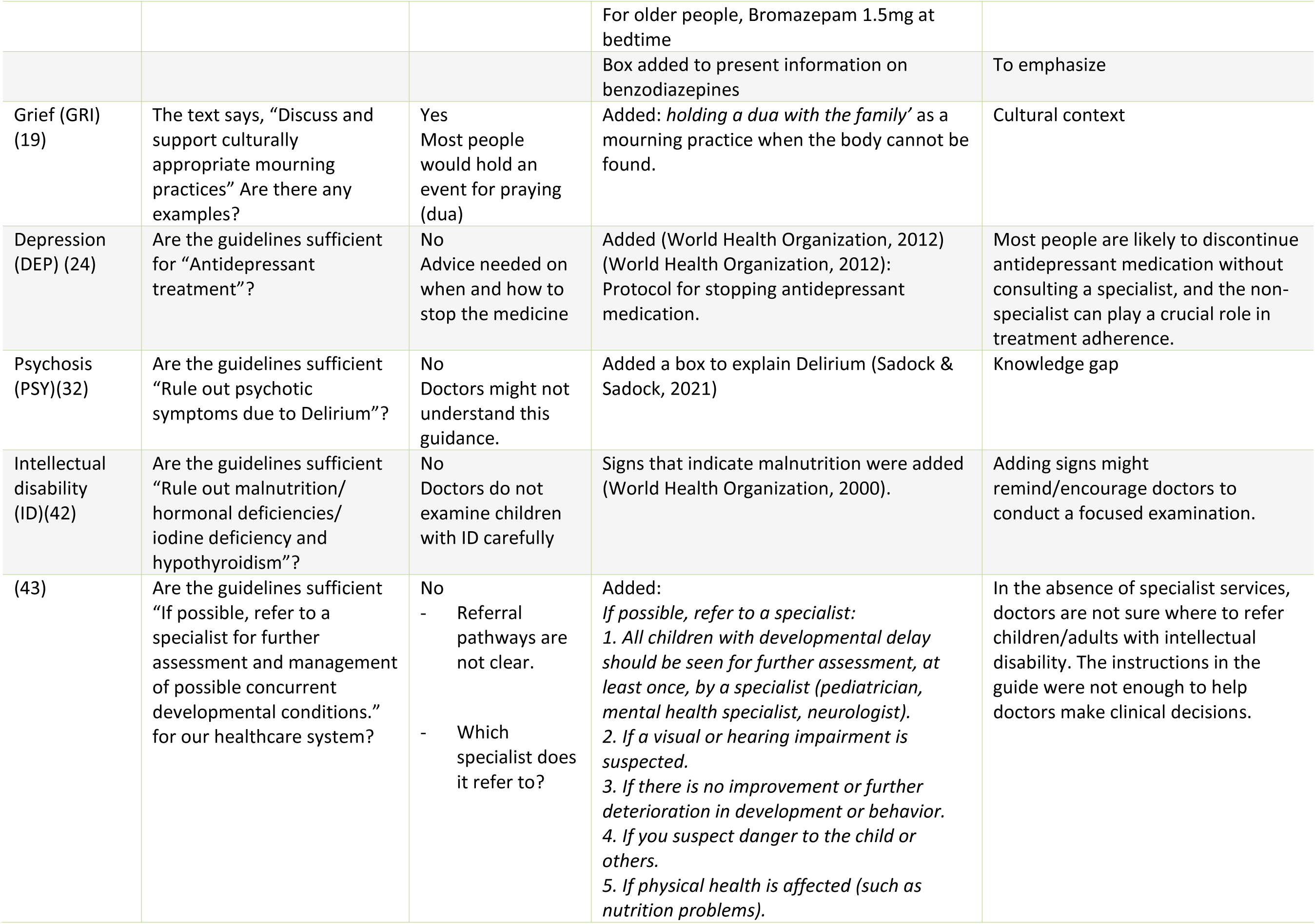
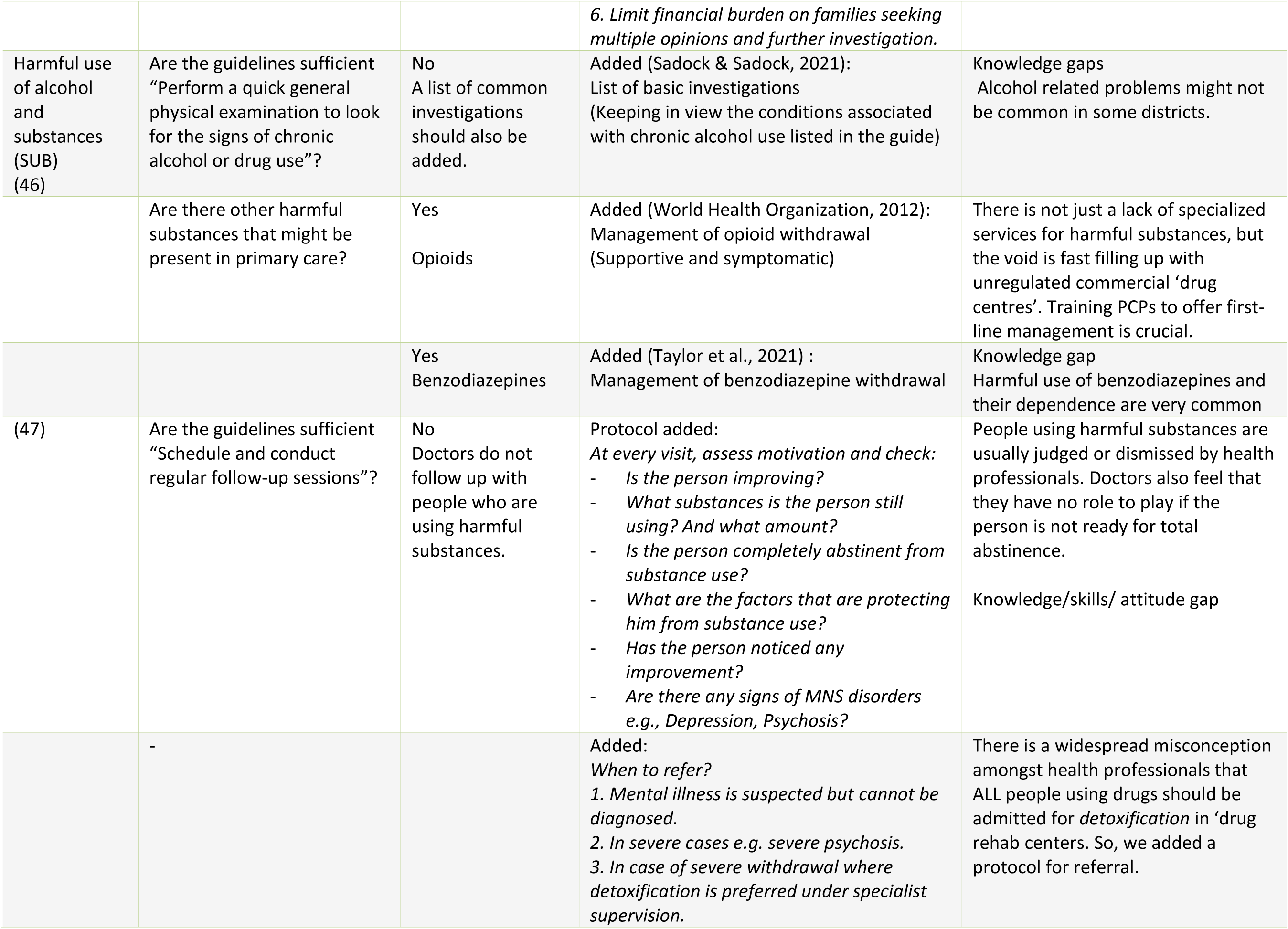
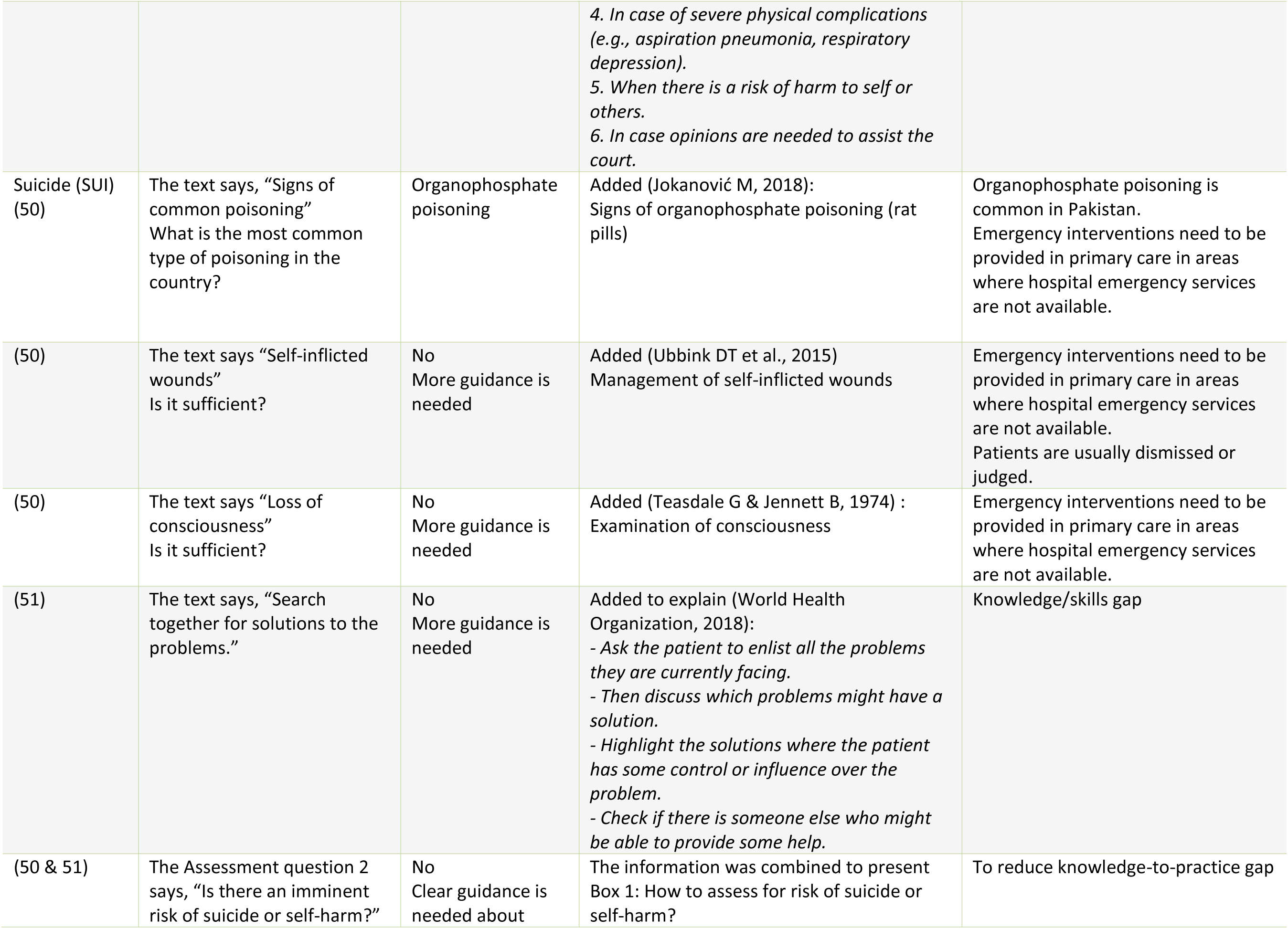
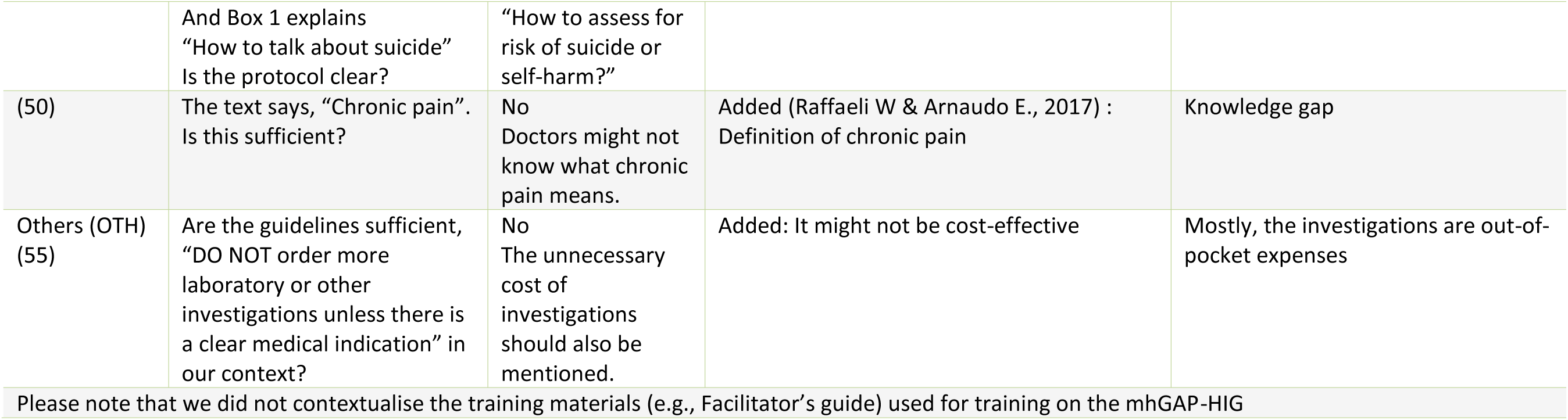
Contextualization of the mhGAP-HIG.

### 2. Contextualizing the mhGAP-HIG to bridge the gaps

We contextualized the mhGAP-HIG to bridge the four identified gaps as follows:

#### 2.1 To address knowledge, skills and attitude gaps

To address this gap, the content of each module was examined based on these guiding questions:

a. Is the content relevant in the healthcare context?
b. Is additional information needed to explain the content?
c. Is there a cultural-specific consideration?
d. Is there a language consideration?

Table 2 presents the identified content associated with the respective challenge; expert recommendations; adapted content; and the rationale for the modification.

#### 2.2 To address knowledge to practice gaps

To address the existing knowledge to practice gaps, the content of each module was examined based on these guiding questions:

a. Is the content applicable in the healthcare context?
b. Is the content applicable in the cultural context?
c. Does the content align with existing resources?
d. Do PCPs have the skill to apply the content?

Even if the content was adapted to the local context, a major challenge identified was a substantial deficiency in the skills of non-specialists for adequate mental health assessment and delivery of psychosocial interventions. It was also noted that the mhGAP-HIG outlines the symptoms but lacks directions on how to elicit them, leaving PCPs to rely on their existing skills. The PCPs also shared their lack of confidence in applying the newly acquired information independently in their clinical practice. We addressed this gap by adding evidence-based interview questions for assessment and instructions to deliver psychosocial interventions in all modules of the guide (World Health Organization, 2019). Finally, all interview questions and instructions were translated into Urdu following the WHO guidelines (World Health Organization, 2010). Some examples of instructions and interview prompts are shown in Table 3.

**Table 3:**
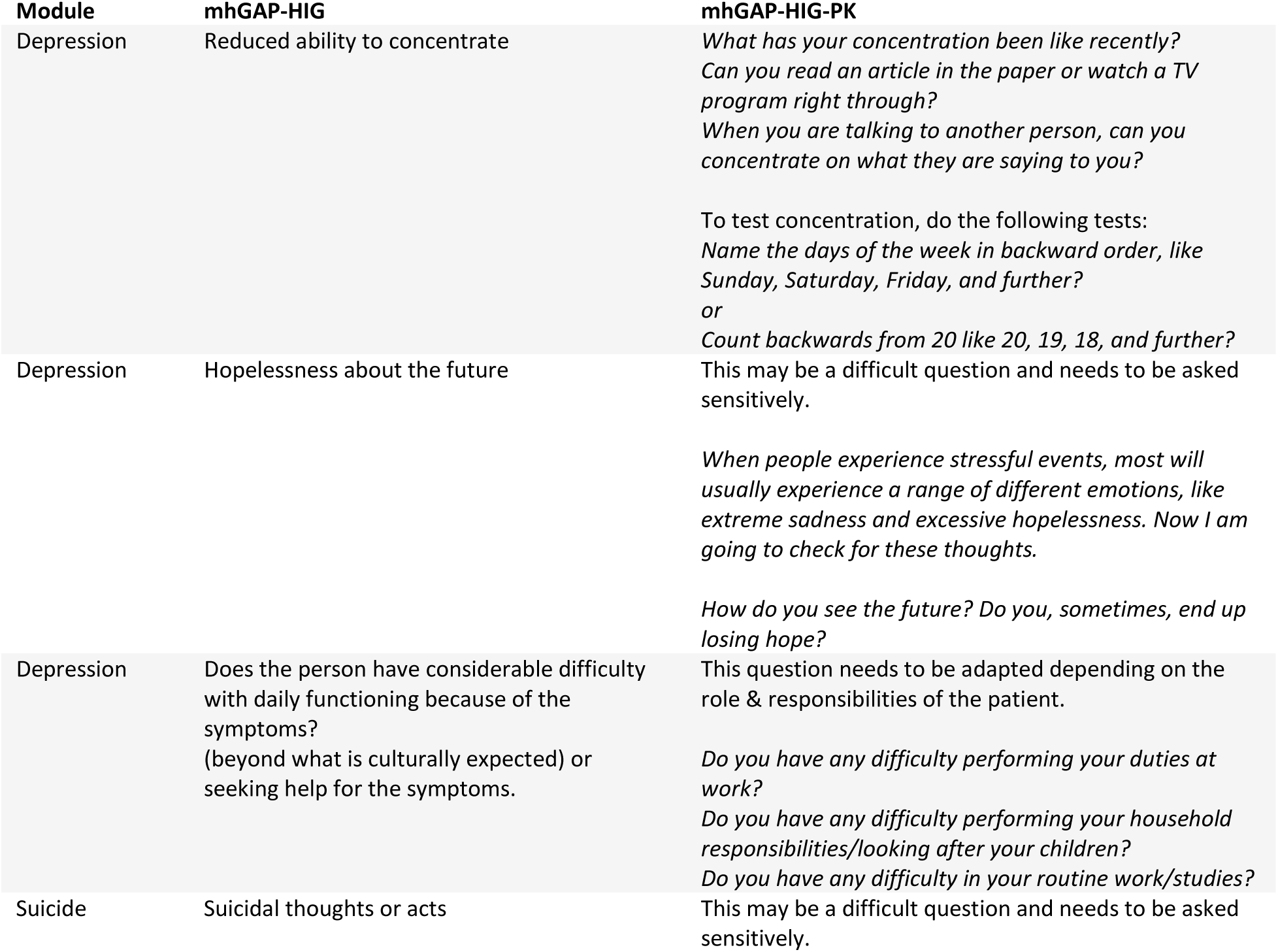
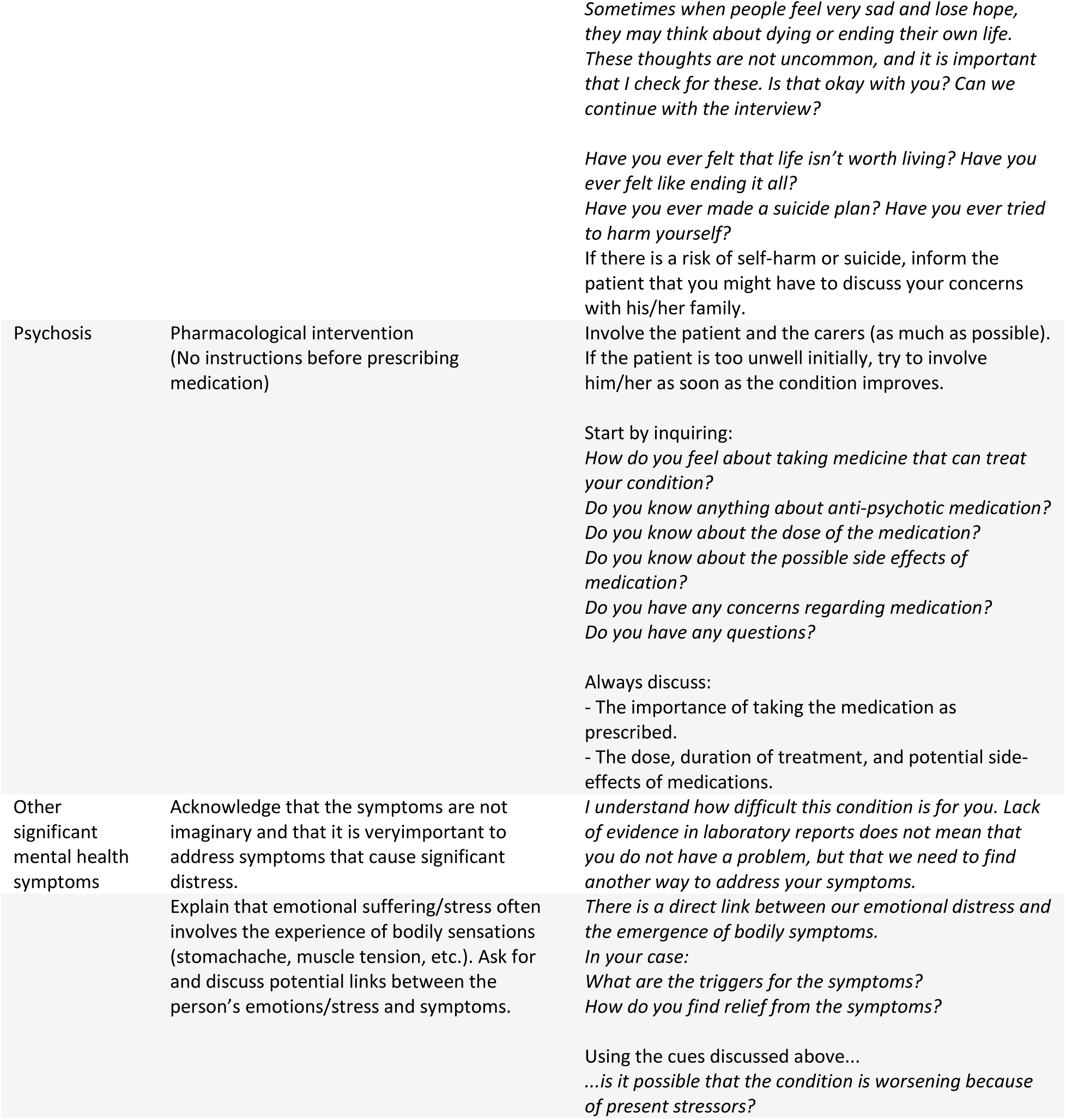
Interview questions for assessment and instructions to deliver psychosocial interventions.

#### 2.3 To address the training resources gaps

Through the FGD and KIIs, two challenges identified for implementing the mhGAP-HIG as a training resource were readability of the guide and its availability. To address readability issues, such as small font size and the dense presentation of the symptoms list and interventions, we performed a comprehensive revision of the guide’s format and visual presentation, focusing on improving clarity and user engagement. For instance, the assessment of depression in mhGAP-HIG, which was previously condensed into a single page, was elaborated upon across five pages in the adapted guide, after incorporating interview questions and technical instructions. To improve user experience, we enhanced navigational clarity by incorporating headings, tables, and boxes e.g. to elucidate conditions like ‘mania’ and ‘prolonged grief disorder’. These adjustments are aimed at enhancing the guide’s effectiveness as a training resource, making sure that essential information is not just easier to access and understand, but also more engaging for the users.

To address the challenge related to accessibility, including the financial implications of printing hard copies on a large scale, we developed a mobile application (mhGAP-HIG-PK) for both iOS and Android platforms (See Figure 1) to serve as a handy reference guide for the primary care staff. During the app development process, the tech team ensured that the design was simple, user-friendly, and multilingual (English and Urdu) to accommodate users with limited tech literacy. Throughout the app development process, trained PCPs were actively involved to give feedback to ensure intuitive navigation and easy access to information, making the app user-friendly and efficient.

**Figure 1.**
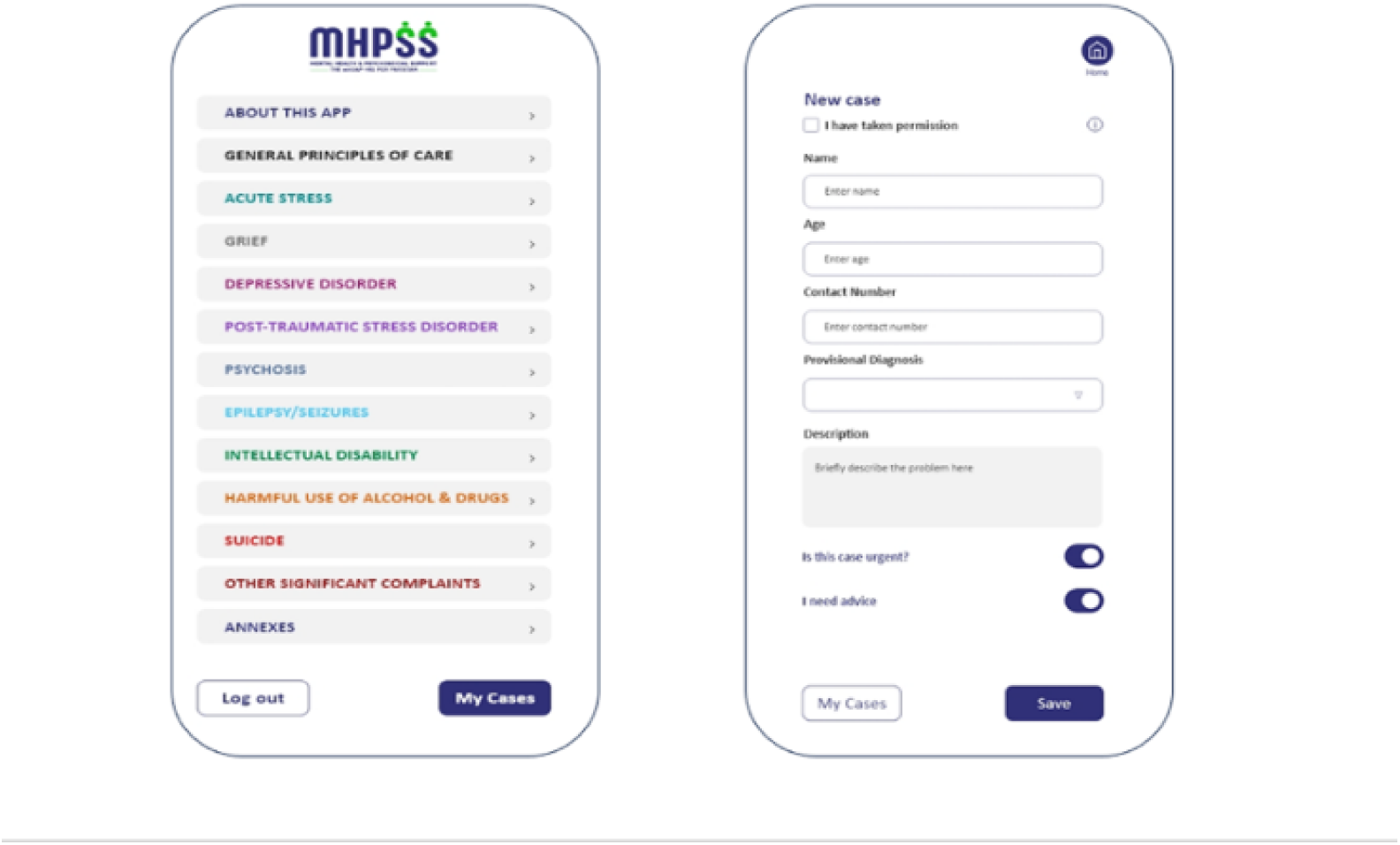
The mhGAP-HIG-PK mobile application: Images of the home page and submission of ‘My cases’.

#### 2.4 To address the supervision gaps

Another critical barrier identified was the lack of a mechanism for supervision due to the severe dearth of supervisors, their unavailability in remote areas, lack of formal referral, and data collection mechanisms. To overcome the supervision gaps, an additional feature was integrated into the mhGAP-HIG-PK mobile application to help PHCWs submit clinical cases to seek supervision. This feature allows the trained PCHWs to send clinical information in real-time to a centralized database^2^ to share with the supervisors. An option to request urgent advice was also added, further supervision process and outcomes are discussed in another paper (Humayun et al., 2025).

### 3. Field testing the training resources

The mhGAP-HIG-PK guide and mobile application were field tested to determine their effectiveness in the context of Pakistan through online training of trainers and training of PHCWs workshops. The pre- and post-training knowledge evaluation of the PHCWs showed a significant improvement of 8.67% (p < 0.05) across all modules.

The qualitative feedback from both the trainers and the PHCWs on the content of the guide was very encouraging. They agreed that the training resources were user-friendly, focused, and highly relevant to their clinical work. The readily accessible protocols (assessment and management) prompted a shift from didactic teaching to an interactive discussion and allowed for practicing role plays. The application of adult principles of learning and a hands-on approach to strengthen skills in role plays was much appreciated. The translated interview questions and examination techniques were highly appreciated. These also helped to standardize the performance of trainees in the role-plays. Both the trainers and PHCWs also felt more comfortable with demonstrating and practicing psychosocial interventions with the help of instructions in Urdu.

The trainers suggested that the duration of five days was not enough to cover all modules and that they felt under pressure to cover the content in the time allocated for each module. The PHCWs also expressed dissatisfaction with the limited time for demonstrating their skills and practicing role-plays. They highlighted the strengths and limitations of online training. Since the training was online, a complete video recording of the training workshop was shared with the participants for future reference. The “poll” feature on Zoom was considered an effective strategy to conduct a true/false quiz after each module.

The PHCWs unanimously endorsed that the mhGAP training should be an essential part of the pre-service training across the country.

## Discussion

This study identified key implementation gaps that informed the process of contextualizing and field testing the mhGAP-HIG for Pakistan. It is the first systematic attempt to adapt the guide in the context of any low-and-middle income country (LMIC) to address the challenges related to practice, training, and supervision. This study is unique because it describes a systematic and culturally grounded approach to contextualize mhGAP-HIG, informed by detailed stakeholders’ input, an in-depth inquiry into existing challenges and hands-on lessons. Through this process, we explored implementation barriers, aligned with the core principles of cultural contextualization and implementation (Faregh et al., 2019).

Furthermore, we utilized a multiphase approach involving both specialists and nonspecialists to refine the contextual components of mental healthcare. Our adaptation process also considered structural realities of Pakistan’s existing healthcare system, such as workforce limitations and scalability challenges, thus it ensures relevance to sustainability. Despite limitations during the training phase, we were successful in testing the complete guide, which provided us with better insights into resource needs, planning, and logistic feasibility for future implementations and scale-up.

We have identified critical gaps that are interrelated and present implementation challenges for the guidelines in the healthcare and cultural contexts of Pakistan. Similar challenges have been iterated with cultural nuances in other LMICs for mhGAP-IG (Faregh et al., 2019a; Petagna et al., 2023a; Siriwardhana et al., 2016; Spagnolo & Lal, 2021). The WHO (2021) has also highlighted similar barriers during pilot implementation of mhGAP-HIG in other LMICs. Based on earlier field experiences in Pakistan, we recognize that the complexity of implementation is rooted in diverse sociocultural and systemic factors. Therefore, it is essential to systematically address these challenges through integration of cultural knowledge into existing healthcare systems, and ethical consideration into both contextualization and implementation efforts (Gómez-Carrillo et al., 2020).

Our study contributes to literature by a comprehensive exploration of these challenges, and contextualization of mhGAP-HIG accordingly. In the light of existing literature, we now discuss some key findings that informed our efforts to contextualize the mhGAP-HIG for Pakistan.

### Knowledge, attitude, and skills gaps

We identified that inadequate pre-service training leads to substantial gaps in knowledge, skills, and attitudes among PCPs in Pakistan. At the level of undergraduate medical training, Psychiatry is not taught as a mandatory subject and is often marginalized. This has also been noted in other countries in our region where outdated curricula and inconsistent quality of teaching across medical institutes result in low standards of training in mental health at undergraduate medical education (Marahatta et al., 2021). A community-based survey in Pakistan revealed that almost two-thirds of the general physicians were unaware of either the diagnostic criteria of depression and anxiety or recommended interventions and instead relied on unscientific practices as prescribing benzodiazepines for mental disorders (Naqvi et al., 2012). In recent years, recommendations to include mhGAP guidelines in pre-service training curricula and an emphasis on inclusion of psychiatry as a compulsory subject in undergraduate medical curriculum have been emerging (Chaulagain et al., 2020; Sagar & Sarkar, 2016; World Health Organization, 2025). .

To address these gaps, we adapted the guide reflecting our healthcare context. For instance, we added information to prevent the malpractice of prescribing benzodiazepines (Naqvi et al., 2012), regulate the usage of antidepressants (Chaudary & Ehtesham, 2023), and the management of opioid withdrawal (Kaswa, 2021; Ochani et al., 2023). Additionally, we included an information box on delirium, recognizing that mere instruction of ruling it out is insufficient. Researchers have observed that delirium is often misdiagnosed and mismanaged, despite its high prevalence among elderly patients in Pakistan (Sabzwari et al., 2014).

It is well established that training PCPs not only contributes to improve their knowledge to identify and manage mental health conditions, but also fosters a positive attitude, reduces stigma and helps overcome their reluctance to diagnose mental health conditions (Ayano et al., 2017; R. C. Keynejad et al., 2022; Salazar et al., 2022). Moreover, a positive change in the attitude of clinicians has been observed following mhGAP-HIG training in other countries, by fostering empathy and tolerance towards people with mental health conditions, regard for their confidentiality and reducing judgmental attitudes (Echeverri et al., 2018).

The gaps in technical knowledge, including an understanding of cultural dynamics of framing symptoms, social stigma, and help-seeking behaviour, are closely linked to gaps in cultural competencies.

Individuals seeking help may attribute their condition to cultural specific beliefs or may commonly present with somatic symptoms (Humayun et al., 2025). A major concern lies with the misinterpretation of these symptoms, due to limited cultural knowledge of MHC, leading to unwarranted medical investigations and pharmacological treatment (Wazir et al., 2023). Given rich cultural and linguistic diversity of Pakistan, it becomes important to understand cultural idioms of mental illness (Kidron & Kirmayer, 2019). Therefore, cultural contextualization of mhGAP guide is crucial to incorporate local expressions of distress and help-seeking patterns to ensure that tools for identification and management are effective for both trainers and trainees alike (Faregh et al., 2019).

We recognize that lack of awareness and societal stigma surrounding mental healthcare restricts access to appropriate treatment and social support in the country (Azam et al., 2019). Furthermore, many families resort to seek help from traditional or spiritual healers instead of accessing formal mental health services (Shah et al., 2019). Literature suggests that integrating culturally informed and community-focused approaches can improve the quality of mental healthcare, patient participation, and clinical outcomes in Pakistan (Muhammad Adil et al., 2025). Moreover, evidence highlights that collaborating with traditional and faith healers can improve pathways to mental healthcare and prevent delays in professional help-seeking, as healers can be a valuable community resource for early identification and may facilitate referrals to specialists (Burns & Tomita, 2015; Spagnolo & Lal, 2021). .

Consistent with this evidence, our adapted guide includes culturally sensitive content aimed at supporting PCPs to identify and address cultural specific barriers to treatment.

### Knowledge to practice gaps

Findings from our FGDs revealed that PCPs show reluctance to incorporate psychosocial interventions in their routine practice because of practical challenges such as excessive workload, short consultation times, and inadequate clinical space. These observations are consistent with prior studies demonstrating how psychosocial aspects of assessment and management received lesser attention (Bitta et al., 2020; Spagnolo et al., 2018). Arora et al., (2016) suggested that PCPs prefer psychosocial interventions that are time-efficient, easy to learn, and do not require multiple visits of patients, emphasizing the need for brief and practical strategies that can easily be practiced in primary care settings. Moreover, clear instructions for psychosocial interventions in simple local language help maximize the impact and enhance the functionality and effectiveness of mhGAP guidelines (Arora et al., 2016; England et al., 2015; Faregh et al., 2019).

Considering these challenges, our efforts were directed to incorporate clear language and local practices into standardized protocols. For instance, to enhance the confidence of PCPs in delivering psychosocial interventions, we incorporated step-by-step instructions, with an example to explain problem-solving technique in the General Principles of Care module (Table 2).

In recent years, patient-centered approaches in mental health systems are being included in standard guidelines, reflecting the principle that “people have the right to receive respectful care that is responsive to their needs and values” (Bird et al., 2022; Larson et al., 2019). Moreover, there is a strong emphasis on the active involvement of service users in both advocacy and service delivery processes (Semrau et al., 2016). Aligned with this approach, we explicitly included structured questions to guide PCPs in engaging patients during assessment and treatment planning.

### Training resources gaps

Addressing training resources gaps requires a focus on readily accessible resources. Evidence from resource-constrained settings suggests that providing the guidelines in hard copy, supplemented with local language text (Momotaz et al., 2019), or through mobile application (Echeverri et al., 2018) has been effective in bridging knowledge to practice gaps. Therefore, we published an adapted, user-friendly guide with instructional content in both Urdu and English. Additionally, to improve accessibility, especially for PCPs working in their busy clinics, we developed a mobile application offering full access to the adapted protocols without the need to carry hard copies.

The use of diverse teaching methods beyond traditional didactic practices are well documented in enhancing learning experience and improving training outcomes. For example, the mhGAP training in Ukraine and South Sudan incorporated strategies such as case-based discussions, experiential learning, and apprenticeship models—pairing less experienced providers with more experienced ones (World Health Organization, 2021). We included role-plays alongside printed training resources that helped strengthen the confidence and skills of participants, as reflected in their feedback.

Although other trainings had set a precedent (World Health Organization, 2021), this was a first attempt to include CPs in mhGAP training in Pakistan. We were unable to explore the implementation challenges related to their practices prior to training, due to logistics constraints. Bird et al. (2022) identified barriers to implement new psychosocial interventions among psychologists, which include the role of personal willingness, inadequate infrastructure and resources. Notably, participants in our training also acknowledged the gaps in their pre-service training and strongly recommended the integration of mhGAP guidelines into standard training curricula. As observed earlier, bringing PCPs and CPs together in a single training cohort not only strengthened the learning experience but also facilitated a shift from a bio-medical to bio-psychosocial approach, and hence improved access to psychosocial interventions through task sharing (Kola et al., 2021).

To address other limitations of resources identified in our FGDs, it is suggested to engage relevant government and non-governmental stakeholders. To minimize providers’ workload and availability of psychotropics, a feedback loop mechanism between PCPs as service providers, and policy makers or donor organizations is essential (Faregh et al., 2019). This approach depicts the importance of active stakeholder engagement, which requires a well-established healthcare system to support effective service delivery.

### Supervision gaps

There is a broad consensus that post-training supervision is essential for the effective implementation of mhGAP guidelines (Al-Uzri et al., 2024; Faregh et al., 2019; Tarannum et al., 2019). Our greatest challenge was to strategize a solution to provide supervision and support for healthcare providers in settings marked by a severe shortage of supervisors, lack of incentives to engage available supervisors, and significant geographical barriers. These challenges are not unique to our context; similar supervisory barriers have been reported in other LMICs (Echeverri et al., 2018; World Health Organization, 2021).

Moreover, we recognized the importance of integrating supervision mechanisms with context-specific data collection systems. Such integration is critical for policy reformation, improving quality care, and guiding strategic scale-up and sustainability (Faregh et al., 2019; R. Keynejad et al., 2021; R. C. Keynejad et al., 2018).

In view of documented resource limitations in LMICs, peer supervision and technology-assisted models are increasingly recommended as effective, scalable, and sustainable approaches to post-training support (Fu et al., 2020; Kemp et al., 2019; Kola et al., 2021). Digital or mobile-based solutions have been successful in supervising non-specialists, particularly in remote community settings with limited access to specialists (Ojagbemi et al., 2022; Pokhrel et al., 2021). Furthermore, to overcome the referral challenges iterated in low-resource settings, particularly in humanitarian contexts where formal referral systems are lacking, remote supervision ensures standardized care (Al-Uzri et al., 2024; Bolton et al., 2023; Echeverri et al., 2018). As part of the broader MHPSS model, the mhGAP-HIG-PK app was developed to support remote supervision, data collection, and establish a referral mechanism, even in areas with limited internet access (MoPDSI, 2022a). When the model is fully implemented, the model envisions the training of PCPs as part of a comprehensive MHPSS service provision, in which the workforce is trained at multiple levels to offer interventions at primary care and to operationalize an effective, digitally supported referral mechanism across services.

At the time of submitting this paper, our contextualized guide had been used to train over a hundred PHCWs in Khyber Pakhtunkhwa (Humayun & Najmussaqib, 2024). They were provided supervision for three months via the mhGAP-HIG-PK mobile application (Humayun et al., 2025). This experience not just validated the feasibility of remote supervision but also demonstrated a positive impact on the performance of PHCWs in delivering mental healthcare.

## Limitations

We acknowledge our study limitations. First, we were able to identify some technical and some system-level barriers, there might be other factors, such as sociopolitical barriers that may impact the health system and affect mental healthcare delivery. Second, we used non-probability sampling, both for the selection of trainers and training participants, due to the excessive workload on healthcare professionals during COVID-19. Future studies may adopt predefined selection criteria, their level of motivation, and willingness to use psychosocial interventions in their routine clinical work.

Third, the field testing of the guide was conducted on a small group of participants, and only knowledge component was evaluated due to time and resource constraints. Future studies may assess knowledge, skills, attitude, and confidence of the participants, both pre- and post-mhGAP training. Finally, we employed an uncontrolled pre-post design, limiting both internal and external validity. Future work may use a more robust design to demonstrate the effectiveness of the adapted mhGAP-HIG.

## Recommendations

Based on our research, we suggest that contextualizing the mhGAP-HIG and conducting related training even with supervision—should not be planned in isolation. We also believe that PCPs should be trained as part of a comprehensive MHPSS service, wherein the workforce is trained at multiple levels to offer interventions at community, primary, and tertiary care levels. We strongly recommend incorporating the adapted guide into the preservice training of healthcare providers in Pakistan, as this would help bridge the knowledge gaps (Iversen et al., 2021; World Health Organization, 2013). To achieve any lasting impact, systematic policy support and continued commitment from both provincial and federal governments, with sustained programmatic backing from donor agencies would remain crucial to overcome these challenges in Pakistan (Petagna et al., 2023; Spagnolo et al., 2020).

There is a need to integrate and strengthen health information systems, and to report data on a core set of mental health indicators (Setoya & Kestel, 2018). Future research should focus on the sustained impact of implementing the guide, including changes in practices, clinical outcomes, and policy-level improvements. Scalability of the MHPSS service across all provinces necessitates active engagement of relevant stakeholders, including government and community-based organizations. Lastly, to ensure effective implementation and sustainability, the healthcare system should be reformed to address key structural challenges, such as excessive workload, inadequate workspace, availability of psychotropics, and human resources.

## Data Availability

The data that support the findings of this study are available from the Health Section at the Ministry of Planning, Development & Special Initiatives, Government of Pakistan. Confidentiality restrictions apply to the availability of the data used under the license for the current study, and so are not publicly available. However, the data can be made available from the authors upon reasonable request after formal approval from the Ministry of Planning, Development & Special Initiatives, Pakistan.

## Acknowledgement

We would like to thank Faisal Rashid Khan and Sarah Nasir for their invaluable contributions towards the process of contextualization of the guide in 2021. We are grateful to Dr. Peter Ventevogel for his support during this project and his comments on an initial draft of this paper.

The final phase of contextualization of the guide and field testing was part of a project which was supported by UNICEF, Pakistan. We also thank International Medical Corps, Pakistan for supporting the initial phase of contextualization in 2020, and the pilot testing of the mhGAP-HIG-PK resources in collaboration with KP Health Department in 2023-2024.

Lastly, we extend our sincere gratitude to the Ministry of Planning, Development & Special Initiatives for endorsing and publishing the guide as a national resource for implementation across the country.

## Ethics approval and consent to participate

This study was conducted as part of the Mental Health and Psychosocial Support Project, approved by the Ministry of Planning, Development & Special Initiatives in compliance with ethical standards and consent protocols under letter no. 6(262) HPC/2020.

## Consent for publication

All authors consented to publication.

## Author’s contributions

AH- Conceptualization, Methodology, Project administration, Supervision, Data curation, formal analysis, Writing-original draft, Writing-review and editing.

NM- Writing-original draft, Data curation

AN- Methodology, formal analysis, Writing-review and editing

IH- Data curation

MA- Project administration

## Abbreviations

mhGAP-IG: mhGAP Intervention Guide;
mhGAP-HIG: mhGAP Humanitarian Intervention Guide;
mhGAP-HIG-PK: mhGAP Humanitarian Intervention Guide for Pakistan;
MHPSS: Mental Health and Psychosocial Support;
MoPD&SI: Ministry of Planning, Development & Special Initiatives;
MoNHSR&C: Ministry of National Health Services, Regulations & Coordination;
WHO: World Health Organization;
iNGOs: International Non-governmental Organizations;
PTSD: Post-Traumatic Stress Disorder;
FGD: Focus Group Discussion;
ToT: Training of Trainers
COVID-19: Corona Virus Disease
LMICs;: Low- and Middle-Income Countries
PCPs: Primary Care Physicians
PHCWs: Primary Health Care Workers
CPs: Clinical Psychologists.

## Footnotes

1 Owing to its pungent smell, Ammonium hydroxide is commonly used in emergency rooms as an aversive treatment for patients with a dissociative state in Pakistan.

2 MHPSS Data Privacy policy has been documented. Moreover, all data is encrypted in transit to avoid its interception and misuse.

